# Utilizing the Amide Proton Transfer Technique to Characterize Diffuse Gliomas Based on the WHO 2021 Classification of CNS Tumors

**DOI:** 10.1101/2023.08.24.23294427

**Authors:** Elena Filimonova, Anton Pashkov, Norayr Borisov, Anton Kalinovsky, Jamil Rzaev

**Affiliations:** FSBI “Federal Center of Neurosurgery”, Novosibirsk, Russia; Department of Neurosurgery, Novosibirsk State Medical University, Novosibirsk, Russia; Department of Data Collection and Processing Systems, Novosibirsk State Technical University, Novosibirsk, Russia; Department of Neuroscience, Institute of Medicine and Psychology, Novosibirsk State University, Novosibirsk, Russia

**Keywords:** brain glioma, amide proton transfer imaging, arterial spin labeling, apparent diffusion coefficient, qualitative MRI features

## Abstract

**Purpose:** Diffuse gliomas present a significant challenge for healthcare systems globally. While brain MRI plays a vital role in diagnosis, prognosis, and treatment monitoring, accurately characterizing gliomas using conventional MRI techniques alone is challenging. In this study, we explored the potential of utilizing the amide proton transfer (APT) technique alone or in combination with other quantitative MRI sequences to predict tumor grade and type based on the WHO 2021 Classification of CNS Tumors.

**Methods:** Forty-two adult patients with histopathologically confirmed brain gliomas were included in the study. They underwent 3T MRI imaging, which involved APT, arterial spin labeling (ASL), and diffusion-weighted imaging sequences. Multinomial and binary logistic regression models were employed to classify patients into clinically relevant groups based on MRI findings and demographic variables.

**Results:** We found that the best model for tumor grade classification included patient age along with APT values. The highest sensitivity (88%) was observed for Grade 4 tumors, while Grade 3 tumors showed the highest specificity (79%). For tumor type classification, our model incorporated four predictors: APT values, necrosis, and the presence of hemorrhage. The glioblastoma group had the highest sensitivity and specificity (87%), whereas balanced accuracy was the lowest for astrocytomas, indicating that the model performs better at detecting patients with glioblastoma rather than astrocytomas.

**Conclusion:** The APT technique shows great potential for noninvasive evaluation of diffuse gliomas. The changes in the classification of gliomas as per the WHO 2021 version of the CNS Tumor Classification did not affect its usefulness in predicting tumor grade or type.

## Introduction

Diffuse glioma is the most common primary brain tumor, accounting for approximately 80% of all primary malignant brain tumors in adults [1]. Overall survival and prognosis for relapse, as well as the provision of adjuvant therapeutic regimens, depend dramatically on both tumor grade and morphological subtype [2]. In addition, it is believed that the molecular profile of the tumor can predict the response to treatment [3], [4]. Thus, an accurate evaluation of the malignant potential of diffuse gliomas is absolutely necessary.

The last release of the WHO Classification of CNS Tumors (2021) is focused primarily on molecular diagnostics, where the main factors for grading and tumor type are the combination of IDH mutation and 1p19q codeletion statuses [5]. However, tumor grading is still carried out through visual analysis [6], which is subjective and not always accurate due to tumor heterogeneity [7]. The evaluation of the Ki-67 proliferation index has been recognized as highly beneficial in achieving this objective [8]. However, it necessitates additional immunohistochemical staining. Consequently, despite morphological analysis being regarded as the definitive method for diagnosing diffuse gliomas, there is variability among observers, including experts [9]. Furthermore, detailed histological analysis with tumor molecular profile evaluation is expensive and requires advanced laboratory techniques, which could be a significant problem in middle- and low-income countries [10].

Brain magnetic resonance imaging (MRI) plays a key role in the diagnosis, presurgical planning, surveillance, and treatment monitoring of gliomas [11]. In recent decades, many attempts have been made to accurately determine the type of diffuse brain glioma based on MRI features [11]–[13] or a combination of MRI and demographic characteristics [14]. In several studies, conventional MRI has been shown to predict both the pathological subtype [15] and even the molecular profile [16] of diffuse gliomas. To unify radiological reports and create a common vocabulary, the VASARI scoring system has been developed for a detailed description of brain gliomas (https://wiki.cancerimagingarchive.net/display/Public/VASARI+Research+Project), and its use has already shown promising results [13], [17]. On the other hand, in recent years, there have been significant advancements in image postprocessing techniques that offer a wealth of additional information extracted from conventional T2-weighted, FLAIR, pre- and postcontrast T1-weighted sequences [18]. These radiomic-based methods showed high precision for the prediction of tumor grade, IDH mutation, and 1p19q codeletion status in adults [12], [13] as well as tumor grade [19] and BRAF mutation status [20] in children. However, the signal changes on most conventional MRI sequences lack biological specificity, which limits the accuracy of noninvasive glioma characterization. Therefore, although conventional MRI is readily accessible and provides crucial anatomical details, accurately distinguishing the type and grade of a tumor solely based on conventional techniques appears to be challenging [11], [12].

The apparent diffusion coefficient (ADC) map is an essential conventional quantitative magnetic resonance sequence that is particularly valuable for analysing diffuse gliomas. It is believed that the restriction of diffusion in this map reflects the level of cellularity within the tumor [21]–[23], and a negative correlation between ADC values and glioma grade has been shown in many previous studies [21], [24]–[26].

The advancements and widespread adoption of advanced MRI sequences in recent decades have enabled clinicians and researchers to gather extensive information about tumor structure and physiology, facilitating noninvasive glioma diagnosis and evaluation of treatment effectiveness. Specifically, perfusion techniques can offer valuable noninvasive insights into the microvasculature of tumors. Several perfusion methods are available and utilized in clinical practice, showing promising results [27], [28]. One of the most popular techniques among them is dynamic susceptibility contrast imaging (DSC), which involves measuring the relative cerebral blood volume (rCBV) [12]. A recently published meta-analysis revealed that this technique can accurately predict tumor grade [28]. Another meta-analysis demonstrated the high accuracy of DSC perfusion in determining both the IDH mutation and 1p19q codeletion statuses in patients with diffuse brain glioma [29]. However, it is worth noting that data acquisition protocols for DSC can be relatively complex and vary significantly across different institutions [30]. Furthermore, this technique is unable to provide definitive values for perfusion parameters, which limits the practical applicability of the acquired data. For example, it becomes challenging to establish a universal threshold in such circumstances. Additionally, this procedure is invasive and necessitates the administration of a contrast agent.

The arterial spin labeling (ASL) technique is another promising alternative to invasive perfusion methods, and its utility has been extensively studied in different fields, including neurooncology [31]. ASL has been shown to be useful in the prediction of glioma grade [32] but much less useful in defining the molecular profiles of gliomas [33]. Moreover, studies suggest a strong correlation between the data obtained from ASL and DSC perfusion techniques [34], [35]. However, it is important to note that ASL can only provide measurements of cerebral blood flow (CBF), whereas CBV values are known to better reflect the microvasculature of the tumor [12].

An additional encouraging MRI technique used for imaging diffuse brain gliomas is amide proton transfer (APT), known for its remarkable biological specificity. It is well known that gliomas have higher protein/peptide contents than normal brain tissue [36]. Therefore, information at the protein level could potentially contribute to earlier diagnosis, more accurate delineation of boundaries, and enhanced tumor characterization [37]. The biophysical basis of APT imaging is the ability to detect mobile proteins [38]. The chemical exchange rate of amide protons with water hydrogens is a crucial parameter in this context, as it enhances the clinical utility of APT [39]. In addition, it is a noninvasive procedure that does not require the administration of any contrast agents.

APT showed better diagnostic performance than conventional MRI [40]. Moreover, it has been discovered that it is just as effective as DSC perfusion [41]. A recently published meta-analysis confirmed the effectiveness of the APT technique in distinguishing between low- and high-grade gliomas, as well as its potential for predicting histopathology noninvasively [38]. However, some authors have reported improved accuracy in grade prediction when combining APT with ADC and ASL values [42]. In addition, several studies [43], [44] have found a correlation between APT values within the glioma and tumor cellularity.

Furthermore, these studies have also shown a relationship between APT values and the extent of diffusion restriction as measured by both ADC and DKI techniques [43], [45].

Beyond that, APT seems to be a predictive factor for IDH mutation status as well [39], [46], [47]. It is important to note that the application of APT is not limited to presurgical glioma diagnosis and can also have implications for predicting overall survival, prognosis of recurrence, and assessing treatment outcomes. This has been demonstrated in multiple recently published articles [47]–[49].

As a reflection of its advantages and benefits for clinical applications, numerous articles focusing on the radiological characteristics of diffuse gliomas have been published over the past years. However, it is worth noting that most of these studies were conducted using the WHO 2016 Classification of CNS Tumors, whereas the recently released WHO 2021 update introduces significant revisions to glioma grading. Interestingly, only a few authors have made an effort to determine the subtype of glioma, despite its clear correlation with patient prognosis. Furthermore, there have been only a limited number of research papers published on the practicality of combining conventional MRI with APT data, as well as on the effectiveness of utilizing different quantitative techniques (such as APT, ADC, and ASL) for assessing diffuse gliomas [42], [50]. Moreover, many studies examining the effectiveness of advanced brain MRI techniques, specifically APT and ASL, in patients with diffuse gliomas have been limited by small sample sizes.

Thus, the main objective of this study was to investigate the usefulness of the APT technique, both alone and in combination with other quantitative MRI sequences (specifically ADC and ASL), in predicting the grade and tumor type according to the WHO 2021 Classification of CNS Tumors for diffuse gliomas. Additionally, we aimed to determine whether incorporating descriptive tumor characteristics could enhance the predictive model based on APT. Furthermore, we sought to assess the ability of APT (alone or combined with other variables) to determine the status of IDH mutations, which greatly influences prognosis. Finally, we were interested in examining the relationship between quantitative MRI data and tumor cellularity measured by Ki-67 levels.

## Methods

### Patients

The subjects in this study were patients with first identified brain gliomas who were surgically treated at our hospital in 2023. Forty-two patients (20 males and 22 females, 22 – 76 years of age) with morphologically proven gliomas (according to WHO 2021 criteria) participated in the study. All patients underwent high-resolution brain MRI before surgery. Detailed information on the patients is provided in Supplementary Table 1. Each patient signed a written informed consent form to participate in the study. The study was carried out according to the Declaration of Helsinki and was approved by the local Ethics Committee of the Federal Center for Neurosurgery, Novosibirsk, Russia (protocol No. 4 dated 02-08-2022).

### Morphology

The surgical samples were assessed using the 2021 WHO classification of CNS tumors [5]. Immunohistochemical staining was conducted to evaluate the IDH1 status in each case. Additionally, FISH analysis was performed to detect 1p19q codeletion. Ki-67 levels were assessed using immunohistochemistry.

### MRI acquisition

MR imaging data were acquired using a 3T system (Ingenia, Philips Healthcare, The Netherlands) equipped with a 16-channel receiver head coil. The MRI protocol included high-resolution T1-WI (before and after contrast injection), T2-WI, FLAIR, SWI, DWI, ASL and APT sequences. Details regarding the acquisition parameters used are shown in Table 1.

### MRI data processing

The quantitative maps of APT, CBF (ASL), and ADC were calculated automatically by the MRI system. In each case, the entire tumor was segmented with ITK-Snap software (version 4.0.0, http://www.itksnap.org) using a semiautomatic classification algorithm. Segmentation was performed based on FLAIR and T2-WI (referring to T1-WI and CE-T1WI) for contrast-negative cases and postcontrast T1-WI (referring to FLAIR, T2-WI, and T1-WI) for contrast-positive cases. The segmentation results were saved as binary masks in NifTI format. The ADC, CBF and APT maps were registered and resampled referring to T1-WI using SPM12 with a normalized mutual information cost function and 4th Degree B-Spline interpolation (http://www.fil.ion.ucl.ac.uk/spm/). Subsequently, the tumor binary mask was moved to the APT, CBF, and CBV maps, and quantitative analysis was performed using Pyradiomics tool (https://aim.hms.harvard.edu/pyradiomics).

The following signal intensity characteristics were extracted from the tumor on the APT, CBF and ADC maps: (1) mean, (2) median, (3) 10th percentile (for ADC), and (4) 90th percentile (for APT and CBF). Absolute values of all quantitative MR parameters were used. An illustration of data processing is shown in Figure 1.

**Figure 1.**
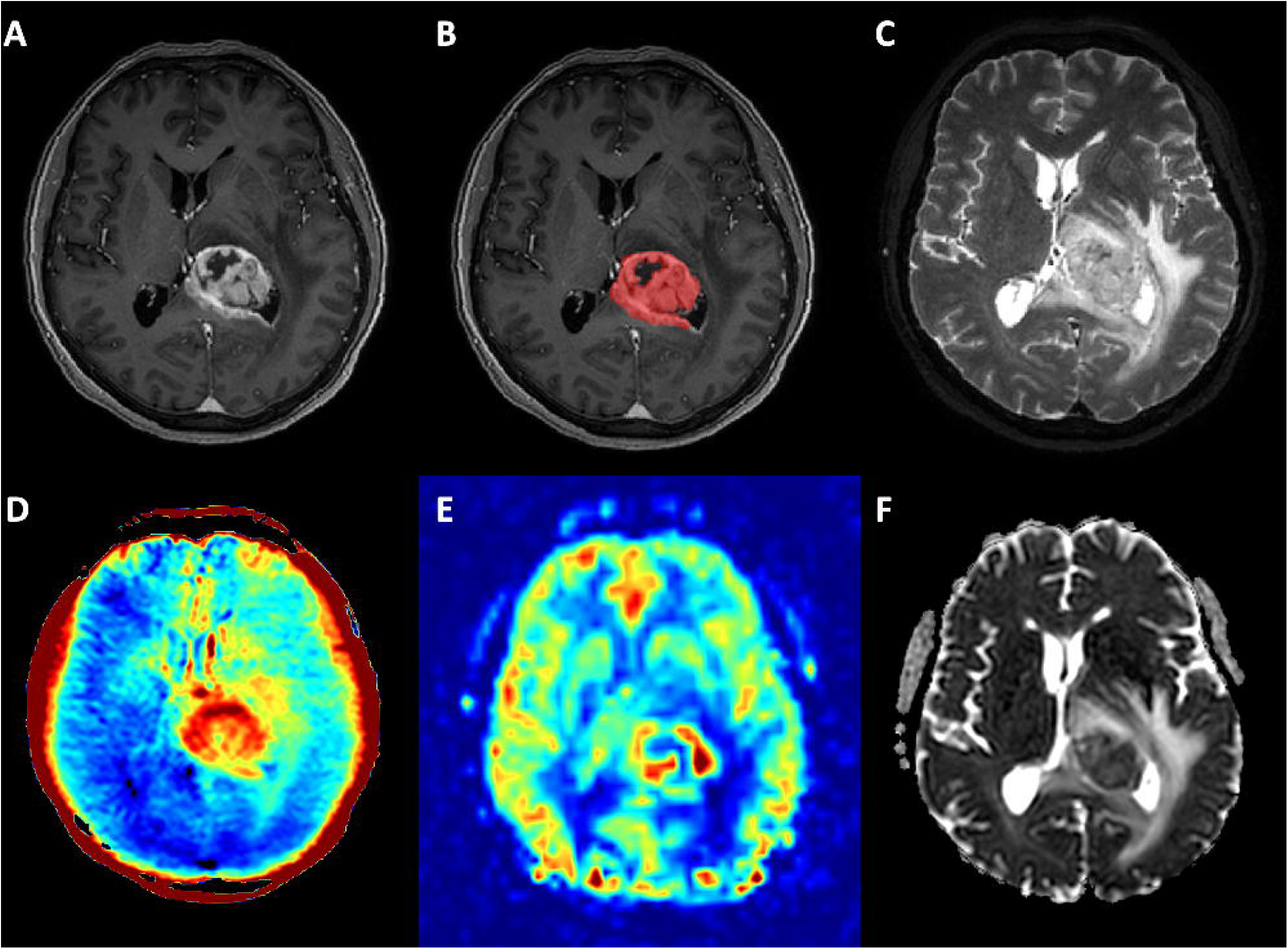
An example of post-processing of MRI data in a patient with glioblastoma (WHO grade 4, IDH1 wild type). A – Axial CE-T1WI shows vivid inhomogeneous enhancement; B – same slice after semiautomatic tumor segmentation; C-axial T2WI; D – APT map demonstrates marked elevation of metabolites in the tumor’s center; E – CBF map (ASL) with slight hyperperfusion from the tumor; F – ADC map shows moderate diffusion restriction.

Two neuroradiologists qualitatively evaluated the MRI data independently (with 5 and 2 years of neuroradiology experience, respectively). Magnetic resonance features were defined according to Visually Accessible Rembrandt Images (VASARI) imaging criteria (https://wiki.cancerimagingarchive.net/display/Public/VASARI+Research+Project), and full results are provided in Supplementary Table 2. Subsequently, qualitative features such as enhancement quality (f4), necrosis proportion (f7), and hemorrhage presence (f16) were evaluated between groups.

### Statistical analysis

Descriptive statistics are presented as the median (interquartile range, IQR), given the relatively small sample size and nonnormal distribution of the data (normality of the distribution was checked with the Shapiro□Wilk test). The relationship between measured variables was assessed with the Spearman correlation coefficient and FDR correction for multiple comparisons. Ninety-five percent confidence intervals are provided in square brackets following the correlation coefficient magnitude. The chi-squared test was used for categorical data analysis. The Mann□Whitney test was utilized for comparison of metric variables grouped by one categorical variable, such as sex, IDH1 mutation and 1p19q codeletion (two levels). Alternatively, in the case of categorical variables such as grade or tumor type, both having three levels, the Kruskal□Wallis test was used instead. A post hoc Dunn test was implemented to estimate the statistical significance between the studied groups in a pairwise manner. To assess the ability of the measured variables to classify patients into the abovementioned clinically meaningful groups (grade, tumor type, IDH1 mutation and 1p19q codeletion), we ran a number of binomial and multinomial logistic regression models. For each model reported in the paper, we provided cross-validated (training/test sample - 70/30) values of accuracy, sensitivity, specificity and area under the curve (AUC). In the case of tumor grade prediction, second grade was used as a reference category. For the tumor type prediction, we chose astrocytoma as a reference. In addition, we used a likelihood ratio (LR) test to assess the difference between distinct versions of nested models, each containing a different number of predictors. Odds ratios and associated 95% confidence intervals were additionally calculated for all predictors included in the final versions of the models. Feature selection was conducted in accordance with the principles of hierarchical regression, where variables that were more likely to be relevant for prediction were initially included in the model based on previous scientific studies, specific investigation objectives, and their interpretability within the overall framework of the study. Before interpreting the resultant models, we sought to ensure that all the logistic regression assumptions held true (i.e., there were no extreme values, outliers or multicollinearity issues in continuous predictors). The values for factors f4, f7, and f16 were obtained through their independent assessment by two neuroradiologists and subsequent calculation of the intraclass correlation coefficient based on a mean-rating (k = 2), absolute-agreement, 2-way mixed-effects model. A p value of 0.05 was considered a threshold for evaluating statistically significant associations. All statistical analyses were run in R (v. 4.3.1, 2023).

## Results

### Patient characteristics

A total of 42 patients (20 males, 22 females) with brain gliomas participated in the study. The median age of the recruited patients was 54.5 years (IQR = 22.75). Male and female patients did not differ in age (U = 267.5, p = 0.24), grade (χ^2^= 3.29, df = 2, p = 0.19) or 1p19q codeletion presence (χ^2^= 0.84, df = 1, p = 0.36). However, they differed in tumor type (χ^2^= 7.21, df = 2, p = 0.027) and IDH1 mutation variables (χ^2^= 5.52, df = 1, p = 0.019). There were 15 patients with glioblastomas, 18 patients with astrocytomas, and 9 patients with oligodendrogliomas. Detailed information on demographic, morphology, and molecular data for each patient is provided in Supplementary Table 1.

### Qualitative MRI features

The full results of the qualitative MRI analysis according to VASARI are demonstrated in Supplementary Table 2. Three characteristics, quality of enhancement (f4), necrosis (f7), and presence of hemorrhage (f16), were chosen for the following analysis. When evaluating the agreement of results obtained from comparing the levels of factors f4, f7 and f16 between two neuroradiologists, we observed intraclass correlation coefficient values of 1 for factors f4 and f7, while factor f16 yielded a value of 0.7, indicating a high degree of consistency in estimates between the raters. As a next step, a chi-square goodness of fit test was performed to determine whether the proportions of these variables were equal between groups of patients with different tumor types and grades. There were significant relations between tumor types and the presence of hemorrhage (χ^2^ = 7.1, df = 2, p = 0.029), necrosis (χ^2^ = 21.4, df = 6, p = 0.0016) and quality of enhancement (χ^2^ = 23.28, df = 4, p = 0.0001). Along the same lines, these variables also differed between patients with different tumor grades (χ^2^ = 8.18, df = 2, p = 0.017 for hemorrhage; χ^2^ = 19.38, df = 6, p = 0.0036 for necrosis; χ^2^ = 21.9, df = 4, p = 0.0002 for enhancement).

### Quantitative MRI features

A Kruskal□Wallis test was performed on the median APT values of the three groups (grades 2, 3 and 4) and revealed that there was a statistically significant difference in these values among the tested groups (χ^2^ = 11.25, df = 2, p = 0.0036). A post hoc Dunn test was further conducted to determine the differences in measured values between each pair of groups. We found the abovementioned differences in grade 2-grade 4 and grade 3-grade 4 pairs (p = 0.03 and p = 0.01, respectively; Figure 2A).

**Figure 2.**
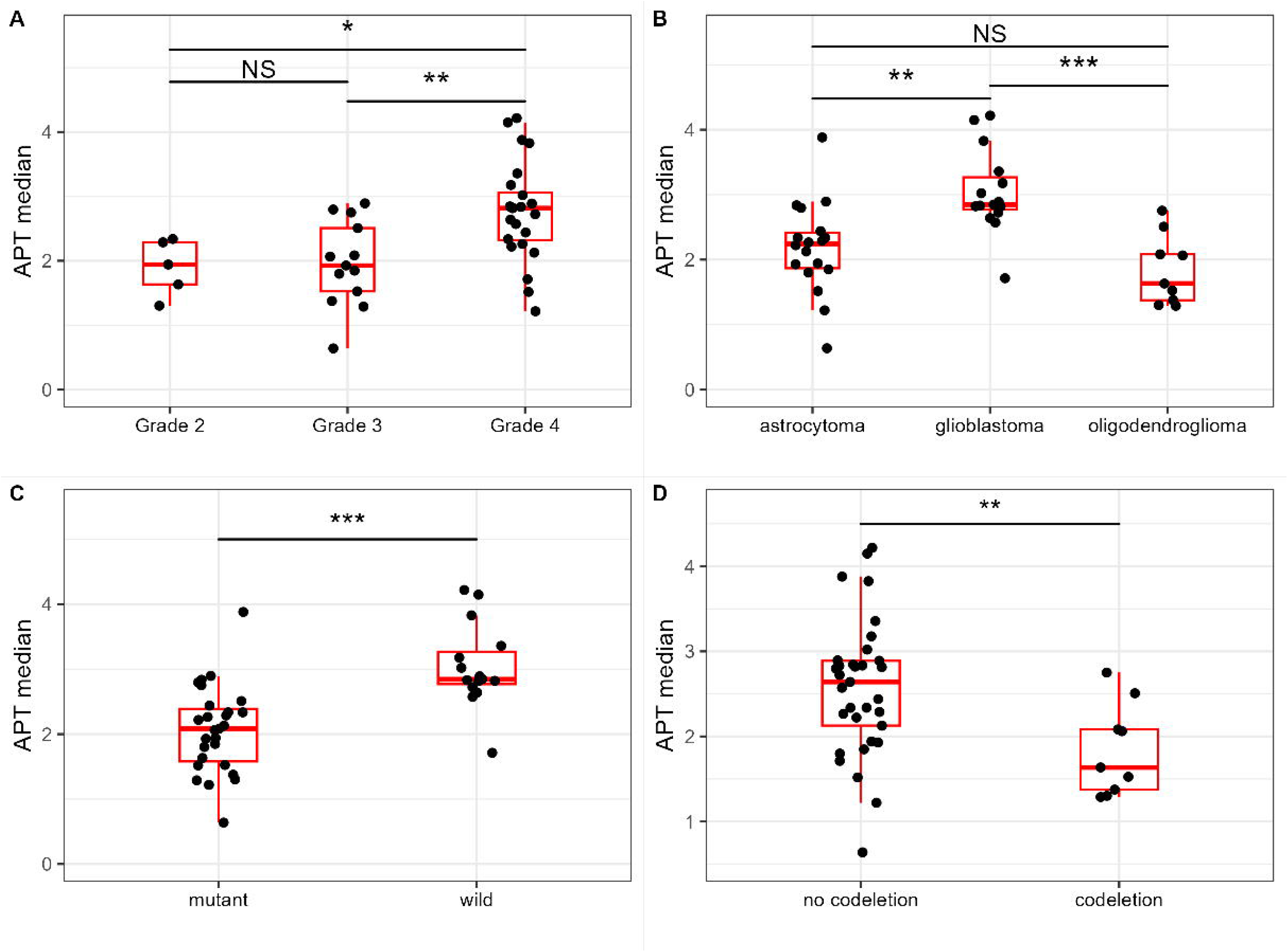
The results of nonparametric statistical comparison of APT median values between groups with Kruskal□Wallis (panels A and B) and Mann□Whitney tests (panels C and D). APT levels are compared among groups using boxplots, where individual data values are represented by black dots and the medians are shown by the thick red horizontal lines. The boxes represent the middle 50% of the data, ranging from the 25th to the 75th percentile. The whiskers extend to 1.5 times the interquartile range. In this figure, NS indicates p > 0.05; * represents p < 0.05; ** p < 0.01; *** p < 0.001.

**Figure 3.**
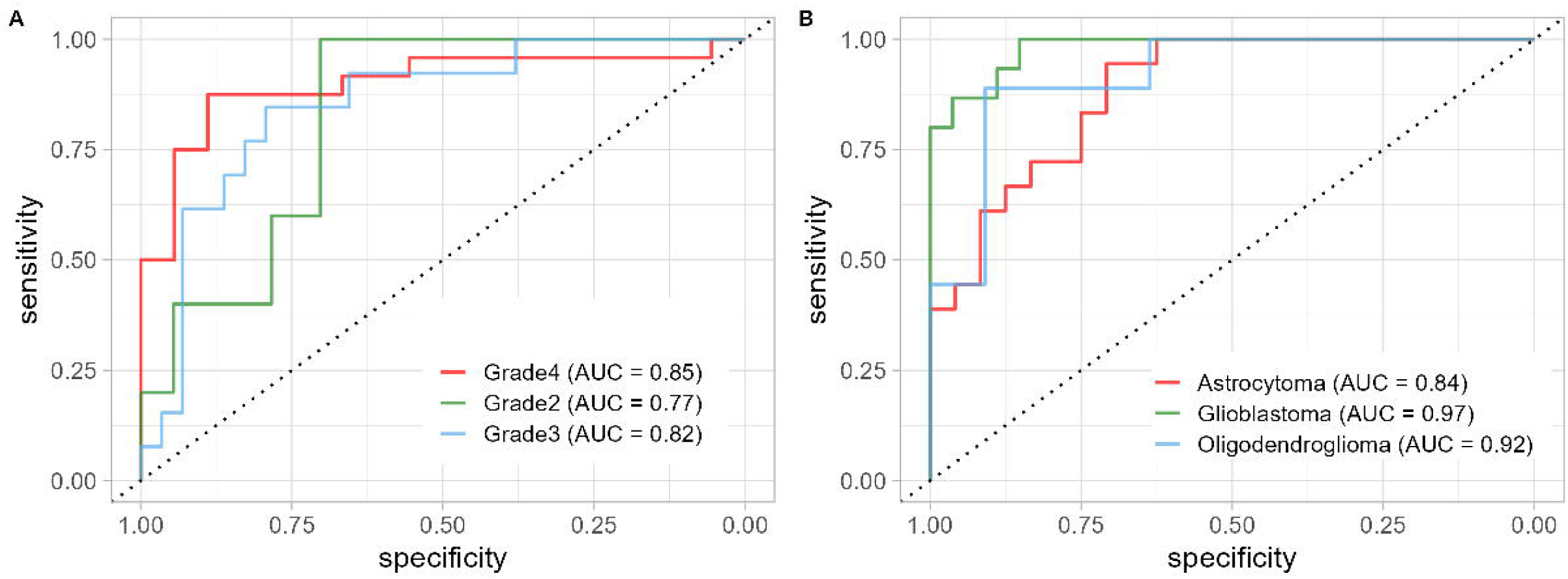
The plot represents areas under the curves (ROC curves) for two multinomial logistic regression models (one-vs-the-rest multiclass strategy) used to classify patients according to their tumor grade (panel A) or tumor type (panel B). Different grades or types are depicted in different colors. The dotted line corresponds to AUC = 0.5 or the random classifier’s performance.

In a similar vein, median APT values were compared between patients belonging to groups with different tumor types, resulting in a rejection of the null hypothesis (χ^2^ = 17.33, df = 2, p = 0.0002). Patients with glioblastoma showed increased median APT values in comparison to those with astrocytoma and oligodendroglioma (p = 0.002 and p = 0.0004, respectively; Figure 2B).

When comparing APT median levels between patients with IDH1 mutant and wild types, the latter group demonstrated increased median values, which were statistically significant (U = 51, p = 7.37*10^−5^; Figure 2C). Similarly, when applied to groups with and without 1p19q codeletion, the Mann□Whitney test indicated the presence of noticeable differences in median APT levels (U = 236, p = 0.0077; Figure 2D). Differences in the mean and 90th percentile APT values were also statistically significant between groups, but these differences were less pronounced than those of median APT. We report these results in detail in Supplementary Table 3.

We did not find any differences in median CBF values between patients with different tumor grades (χ^2^ = 2.6, p = 0.27) or types (χ^2^ = 3.96, p = 0.14). Similarly, these values were not significantly different for patients within the IDH1 mutation (mutant/wild types) and 1p19q codeletion (yes/no) groups (U = 127, p = 0.09 and U = 134.5, p = 0.78, respectively). Comparison of the mean and 90th percentile CBF values showed the same trend and were not reported in the main text of the article.

Likewise, we did not observe any significant differences in median or mean and 10th percentile ADC levels among different grades, tumor types and IDH1 status. Here, we briefly presented only results regarding median ADC values with corresponding data for mean and 10th percentile values provided in Supplementary Table 3. Notably, the Kruskal□Wallis test returned statistically significant differences between tumor grade groups, showing that the null hypothesis about equality of group’s median values can be rejected (χ^2^ = 6.86, df = 2, p = 0.03). However, post hoc Dunn test results revealed no significant differences upon pairwise comparison (p = 0.47 for the grade 2-grade 3 pair; p = 0.09 for the grade 2-grade 4 pair and p = 0.07 for the grade 3-grade 4 pair). Median values for tumor type groups did not differ from each other (χ^2^ = 2.28, df = 2, p = 0.32). The Mann□Whitney test applied to ADC data also did not result in any discrepancies between mutant and wild types (U = 260, p = 0.135).

### Tumor grade and type prediction

To predict a patient’s tumor grade based on a linear combination of MRI parameters as well as clinical and demographic variables, we ran several multinomial logistic regression models, characterized by different numbers of predictors or independent variables. The LR test showed that comparison of the null model and the updated version with the addition of APT values returned statistically significant differences between models (χ^2^ = 13.17, df = 2, p = 0.0014). Including the age variable in the model resulted in further improvement in its performance and led to differences from the model that used APT values as a single predictor (χ^2^ = 8.7, df = 2, p = 0.012). Entering additional variables, such as necrosis, hemorrhage or quality of enhancement, into the model did not result in any significant model improvement, so we did not report them in the article. The best model containing APT and age as the main predictors had a multiclass AUC of 0.82 and an accuracy of 0.71. The balanced accuracy values were 0.5 for grade 2, 0.74 for grade 3 and 0.77 for grade 4. Notably, grade 2 showed a sensitivity of 0% but a specificity of 100%, most likely resulting from the small group size (n = 5). Grade 4 demonstrated the highest sensitivity (88%) among all groups tested while having a moderate magnitude of specificity (67%). Finally, the model was able to correctly classify patients as belonging to grade 3 in 69% of cases, with a specificity value reaching 79%. Table 2 shows the logs of the odds ratio for each predictor included in the model, 95% confidence interval and corresponding p value. In this case, it is evident that the sole significant predictor for this model is the median APT value in the fourth-grade group, which reflects the change in the odds of membership in the target group for a one-unit increase in the predictor. It is also important to highlight that the choice of the second grade as a reference category in our study significantly influences the results presented in the table in such a way that selecting either the third or fourth grade as a reference would yield different sets of model parameters.

Similarly, with the objective of classifying patients into distinct tumor types, we constructed multiple multinomial logistic regression models with varying numbers of predictors. Initially, we included APT median values as a single predictor, following prior grade prediction models, and found that this model exhibited superior performance compared to the null model (χ^2^ = 19.5, df = 2, p = 5.77*10^−5^). Subsequently, incorporating age as a second predictor further improved the model’s performance significantly when compared to the single-predictor model (χ^2^ = 12.85, df = 2, p = 0.0016). Adding the variables “hemorrhage” or “enhancement” as predictors did not result in statistically significant differences from the model with two predictors but demonstrated results trending towards statistical significance (χ^2^ = 5.46, df = 2, p = 0.065 and χ^2^ = 9.3, df = 4, p = 0.054, respectively). The same results were obtained for the necrosis variable (χ^2^ = 9.53, df = 6, p = 0.15). However, it should be noted that a four-predictor model (APT, age, hemorrhage, and necrosis) significantly differed from the two-predictor model (APT and age) and led to improvement in classification performance (χ^2^ = 22, df = 8, p = 0.005; accuracy = 0.74, mcAUC = 0.93). Therefore, it was decided to proceed with this four-predictor model including necrosis and hemorrhage as third and fourth predictors. This final version of the model achieved sensitivity values ranging from 67% for the oligodendroglioma and astrocytoma groups to 87% for the glioblastoma group. The highest specificity score was observed in glioblastoma patients (93%), whereas the lowest specificity score was found in the astrocytoma class (79%) (the oligodendroglioma group had a value of 88%). The balanced accuracy magnitudes were 0.73 for astrocytoma, 0.77 for oligodendroglioma and 0.9 for glioblastoma. The odds ratio for each predictor in the model can be found in Table 3.

### IDH1 status prediction

We further sought to predict the patient’s IDH1 class, that is, whether the patient had mutant or wild-type IDH1. For this end, several binary logistic regression models were developed. As were the cases with tumor grade and type classification, we started with APT median values being the only predictor in the model and found that this model was significantly different from the null model (χ^2^ = 17.62, df = 1, p = 2.7*10^−5^). The inclusion of the age variable resulted in a substantial enhancement in the performance of the model (χ^2^= 12.56, df = 1, p = 0.0004). In total, these two variables provided a model with an accuracy of 0.79 [0.4, 0.97] and an area under the curve of 0.95; however, this model exhibited a low specificity rate of 30%. The likelihood ratio test indicated that further addition of variables to the model did not result in statistically significant differences compared to the model with only two variables. However, the inclusion of the predictor “necrosis” led to an increase in model accuracy up to 1 [0.66, 1], while there was minimal change in the area under the curve, which reached a value of 0.96 (χ^2^ = 5.16, df = 3, p = 0.16). In Figure 4, we present the ROC curves for each of the aforementioned models. In addition, an increase in the median APT and age values by one unit led to the elevation of the odds ratio of membership in the wild-type group (see Table 4 for specific values).

**Figure 4.**
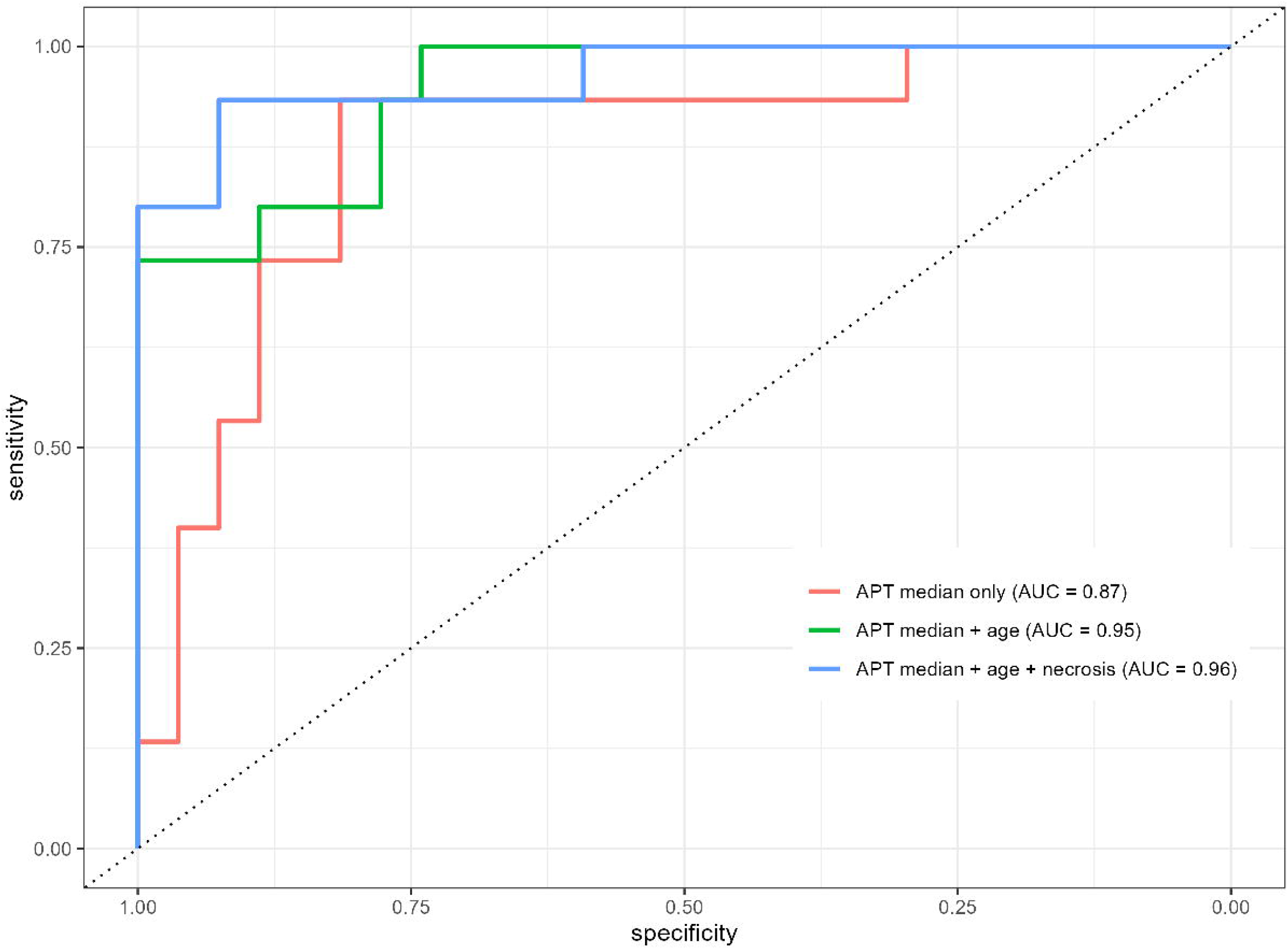
The plot displays areas under the curves for three binary logistic models (IDH1 group prediction) with different numbers of predictors, depicted in different colors. The dotted line corresponds to AUC = 0.5 or the random classifier’s performance.

### Correlations among APT, ADC and CBF values within tumor

We observed a negative correlation between the 90^th^ percentile APT and 10^th^ percentile ADC values (r = - 0.44 [−0.66, −0.15], adjusted p = 0.008; Figure 5). Similarly, there was a weak negative correlation between the mean/median APT values and the 10^th^ percentile ADC values (r = - 0.39 [−0.63, −0.09], adjusted p = 0.016; not shown). No associations were revealed between CBF values (mean, median, 90th percentile) and APT values (mean, median, 90th percentile), as well as ADC values (mean, median, 10th percentile) - p > 0.1 in all comparisons, not shown.

**Figure 5.**
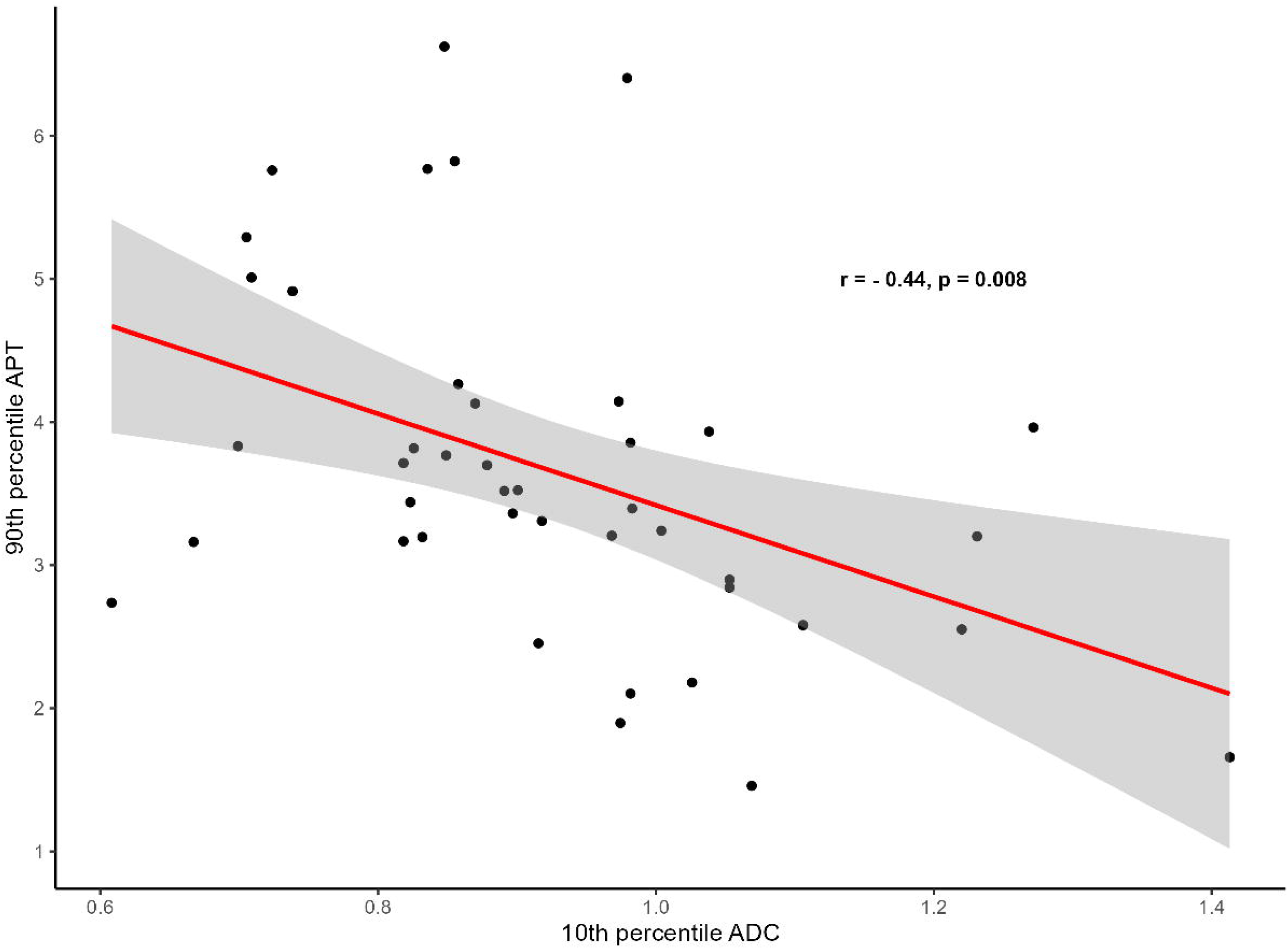
Spearman correlation between 90th percentile APT and 10th percentile ADC values. The gray area around the red line represents the 95% confidence interval. Individual data points are displayed as black dots.

### Correlation between Ki-67 levels and MRI parameters

We found a moderate positive correlation between Ki-67 and the 90th percentile APT values (r = 0.53 [0.26, 0.72], p = 0.003; Figure 6A) and a moderate negative association between Ki-67 and the 10th percentile ADC values (r = - 0.5 [−0.70, −0.22], p = 0.004; Figure 6B). Additionally, mean and median APT values also exhibited significant correlations with Ki-67 levels, although to a lesser extent than the 90th percentile values (r = 0.47 [0.18, 0.68], p = 0.004 for both variables, not shown). Likewise, the mean and median ADC values showed a significant correlation with Ki-67 levels (r = - 0.33 [−0.59, −0.02], p = 0.046 and r = - 0.39 [−0.63, −0.09], p = 0.019, respectively; not shown). No significant correlations were observed between Ki-67 levels and mean, median, or 90th percentile CBF values (p > 0.1, not shown).

**Figure 6.**
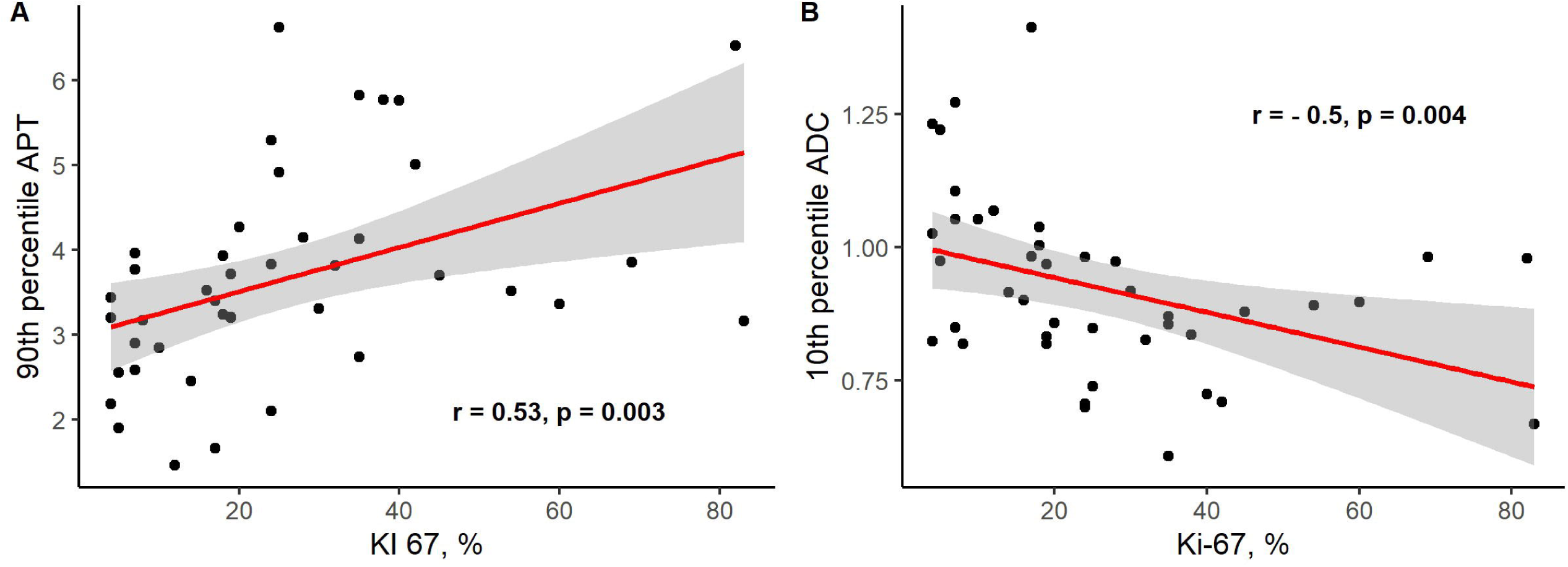
Panel A displays the Spearman correlation between the 90th percentile APT and Ki-67 values. Panel B shows the Spearman correlation between the 10th percentile ADC values and Ki-67 levels in the patients. The gray area around the red line represents the 95% confidence interval. Individual data points are displayed as black dots.

## Discussion

Diffuse gliomas present a major challenge to the modern healthcare system worldwide, leading to a substantial reduction in patients’ quality of life, posing an immediate threat to their lives, and requiring significant direct and indirect financial expenses associated with the prevention, diagnosis, and treatment of such tumors. Brain MRI is essential for diagnosing, planning surgery, monitoring progress, and treating gliomas. However, accurately distinguishing the type and grade of a tumor solely based on conventional MRI techniques seems challenging due to the lack of biological specificity in signal changes, limiting noninvasive glioma characterization. In this study, we explored the potential of using the APT technique, both on its own and in conjunction with other quantitative MRI sequences (specifically ADC and ASL), to predict the grade and tumor type based on the WHO 2021 Classification of CNS Tumors for diffuse gliomas. We discovered that incorporating APT values into the models resulted in a substantial enhancement of their performance. Grade prediction metrics based on median APT values and patient age were the highest for grade 4, with grade 3 and grade 2 demonstrating moderate classification accuracy (all AUC values exceeded 0.75). These findings align well with the literature, which demonstrates the predictive potential of APT levels in distinguishing between low- and high-grade gliomas [38]. Furthermore, we expanded upon these findings by demonstrating the capability of APT to distinguish patients within the high-grade class, specifically those categorized as grade 3 and grade 4. It is crucial to consider this aspect, as there are significant disparities in both overall and median survival rates observed among patients diagnosed with grade 3 and grade 4 diffuse brain gliomas [51]–[53]. Similar results have been reported previously by Guo and colleagues in their study involving 62 patients, further validating our findings [46]. However, the most notable contrast between their findings and ours was the existence of statistically significant differences between grade 2 and grade 3 patients in their study, a pattern that was absent in our data. This discrepancy could be attributed to the relatively small number of patients included in the grade 2 group within our study. We were also able to identify the differences in APT levels among distinct tumor type groups. The most pronounced differences were found between the glioblastoma and oligodendroglioma groups, followed by astrocytoma and glioblastoma. In contrast, there were no statistically significant differences observed between the astrocytoma and oligodendroglioma groups. To our knowledge, these results have not been showcased in earlier studies and provide definitive clinical relevance.

IDH mutation and 1p19q codeletion statuses in patients with diffuse gliomas are well-established prognostic markers [5]. We discovered that patients with wild-type IDH1 had higher APT levels than those with mutant IDH1. Consistent with our current findings, several previously published studies have also shown a similar trend, indicating that elevated APT values can serve as a significant predictor of poor overall survival [47], [54]. In the same vein, it has also been revealed that APT imaging exhibited superior performance over DKI in IDH mutation status prediction [45].

The two 1p19q codeletion groups in our study were also different from each other in terms of APT levels. Specifically, patients without codeletion were characterized by higher APT median values in comparison to patients with codeletion. However, contrary to what we have observed, Su and colleagues in their recent paper failed to identify differences between the groups in a sample of 113 patients with diffuse glioma [55]. Further research is required to address this question, as the presence of 1p19q codeletion classifies the tumor as an oligodendroglioma [5], which significantly improves prognosis for the patient.

Taken together, our findings clearly demonstrate the significant utility of the APT technique in predicting tumor grade and type, as well as identifying IDH mutation and 1p19q codeletion statuses, in patients with diffuse brain gliomas.

Hereafter, a number of multinomial logistic regression models were developed to predict patient tumor types and grades based on APT values complemented by additional parameters used as predictors. The model with the best classification performance for tumor grade, in addition to APT values, included the variable of patient age. This model demonstrated an accuracy of 0.71 and an area under the curve of 0.82, which is considered good classifier performance. The highest sensitivity values (88%) were obtained for Grade 4, while specificity was highest for Grade 3 (79%). Age is a known predictive factor for tumor malignancy and clearly impacts prognosis in patients with diffuse gliomas [56]–[58]. Therefore, entering the age of the patients in the APT-based prediction model looks logical, especially considering that this information is easy to obtain. However, we hypothesize that classification accuracy could be further improved by incorporating parameters not considered in this study.

In the case of tumor type classification, we settled on a model that includes four predictors. In addition to APT values, factors such as necrosis and the presence of hemorrhage were added. This allowed us to achieve an accuracy of 0.74 and an area under the curve of 0.93. Sensitivity and specificity reached their highest values for the glioblastoma group (87%). The balanced accuracy was lowest for the astrocytoma group, which collectively demonstrates the model’s ability to correctly detect patients with glioblastoma most effectively, while performing less accurately for astrocytomas. Conventional MRI is the diagnostic standard for patients with diffuse glioma and provides a detailed anatomical picture, as well as important qualitative tumor characteristics, such as contrast enhancement quality and the presence of necrosis or hemorrhage. Conventional brain MRI has been shown to be useful in predicting the pathological subtype [15] and even the molecular profile [16] of diffuse gliomas. The VASARI scoring system enables the quantification of tumor characteristics, making it easier to compare them between different groups [13].

In general, we found that including additional parameters (specifically the patient’s age, proportion of tumor necrosis, and presence of hemorrhage) into the regression model significantly enhanced the accuracy of predicting type, grade, and IDH mutation status beyond just APT values.

Surprisingly, we did not find differences in ADC values between tumor grades and histological subtypes. Furthermore, there were no significant differences between the IDH mutant and the IDH wild-type groups. These findings are generally inconsistent with previously published articles that revealed the high utility of ADC in the evaluation of gliomas [21], [24]–[26]. The reason could be that the primary purpose of applying ADC is to distinguish between low-grade and high-grade gliomas [24], [25], whereas our sample consisted mainly of high-grade tumors (grades 3 and 4). Another potential explanation lies in our chosen analysis method, wherein we employed an entire tumor binary mask for value extraction. This approach may have inadvertently obscured differences attributable to the inherent tissue heterogeneity of gliomas. Furthermore, it is widely acknowledged that ADC maps derived from higher b-factors offer enhanced diagnostic utility [21], whereas our study utilized the conventional acquisition method with a b-value of 1000 s/mm^2^. It should be emphasized that certain published studies have failed to identify discernible discrepancies in ADC values within tumors when comparing low-grade and high-grade gliomas [59].

Similarly, our study found no significant differences in ASL values among different tumor grades, types, and IDH mutation statuses. These findings are also inconsistent with previous reports in the literature. Previous studies have demonstrated the utility of ASL in predicting glioma grade [34] and assessing IDH status [60]. However, Kang *et al*. did not observe significant differences in the 90th percentile regional cerebral blood flow (rCBF) values between low- and high-grade glioma groups [42]. A recent meta-analysis indicated that ASL shows high accuracy in glioma grading, particularly when considering maximal relative CBF values [32]. It is worth noting that our study employed absolute CBF values, which may explain the discrepancy observed compared to previously published data. Another potential explanation could be attributed to changes made by the WHO’s 2021 Classification of CNS Tumors in grading gliomas, whereas most prior research was based on the WHO 2016 version.

Another noteworthy observation was that the inclusion of ADC or ASL variables in our APT-based regression model did not yield any statistically significant enhancement to the model. Nevertheless, prior investigations have demonstrated that incorporating ADC values into APT may enhance glioma grading [42], [50]. Consequently, further studies are warranted to elucidate this aspect.

We found positive associations between tumor proliferation molecular marker (Ki-67) levels and APT values, as well as negative associations between Ki-67 and ADC values. These results are in agreement with previously published data. In a study by Yin *et al*. (2012), Ki-67 expression was inversely associated with ADC values in tumor parenchyma [61]. Furthermore, Yao *et al*. (2023) demonstrated that minimal ADC values were diagnostically comparable to the Ki-67 proliferation index in assessing pediatric glioma grade [23]. Several prior reports have also shown associations between APT values and Ki-67 levels [62]–[64]. These results support the notion that active tumor cell proliferation is linked to elevated concentrations of mobile proteins within the tumor, leading to an increased APT signal, as well as high cellularity resulting in decreased ADC values. Consequently, both ADC and APT measurements can indirectly reflect the proliferation index and subsequent malignancy of gliomas.

Moreover, our study unveiled a negative correlation between APT and ADC values within the tumor, thus corroborating earlier findings reported in the literature. A negative association has recently been shown between ADC and APT values within the tumor [43]. However, our results indicate that APT exhibits superior diagnostic accuracy when compared to ADC. Therefore, while both APT and ADC values provide insights into tumor cellularity, only APT demonstrates precise prognostic capabilities for determining tumor grade and type.

A previous study highlighted the APT technique as a viable alternative to contrast injection-dependent sequences such as DSC perfusion [41]. However, our findings indicate that APT alone lacks the necessary precision to accurately predict tumor type, and the inclusion of necrosis proportion, which can only be identified through CE-T1WI, is needed. Exploring the potential utility of combining APT and DSC values in presurgical glioma definition represents a promising research direction warranting further investigation. Our study results reaffirm the significant potential of the APT technique for the noninvasive diagnosis of diffuse gliomas. Therefore, it should be routinely incorporated alongside morphological analysis. Another valuable application of APT lies in selecting optimal sites for presurgical stereotactic biopsies. It is well established that targeted biopsy of the most malignant region of a tumor enhances diagnostic accuracy, and this region can potentially be identified using APT maps. It is important to note that beyond its role in presurgical glioma evaluation, APT holds considerable utility in assessing treatment effects and overall prognosis [47]–[49], although these aspects extend beyond the scope of our current study.

This study has several limitations. First, the sample size was small, consisting of only 42 patients, with only five cases presenting low-grade tumors. Second, our brain MRI protocol lacked a DSC perfusion sequence, thus impeding any comparison between the effectiveness of APT and rCBV values and hindering an assessment of their combined utility. Furthermore, there is a lack of follow-up data for the patients who participated in this study.

In addition, within the scope of this study, we also decided to utilize multinomial logistic regression models as classifiers. In doing so, we employed a hierarchical regression approach for variable selection and inclusion in the model. We are fully aware of alternative classifier options that could have been used in this study, such as support vector machines or decision tree classifiers. Similarly, forced entry, stepwise or all-subsets methods could have served as alternatives to hierarchical regression. Each of these methods has its own merits and limitations and cannot be deemed superior a priori over others. Therefore, the choice of methods and approaches was determined by specific research objectives.

In conclusion, the amide proton transfer technique shows significant potential for noninvasive evaluation of diffuse gliomas. To the best of our knowledge, this study represents the initial endeavor to evaluate the effectiveness of APT in the preoperative assessment of gliomas using the WHO 2021 Classification of CNS Tumors. Furthermore, it is a pioneering attempt to integrate APT data with qualitative tumor characteristics and patient demographics to enhance diagnostic accuracy rates. However, given the pilot nature of the study, further studies with larger sample sizes and comprehensive follow-up are clearly necessary to strengthen these findings.

## Data availability

Raw data were generated at Federal Neurosurgical Center Novosibirsk. Derived data supporting the findings of this study are available from the corresponding author on request.

## Abbreviations

ADC: apparent diffusion coefficient
APT: amide proton transfer
ASL: arterial spine labeling
CBF: cerebral blood flow
CBV: cerebral blood volume
CNS: central nervous system
DSC: dynamic susceptibility contrast
IDH: isocitrate dehydrogenase
MRI: magnetic resonance imaging
WHO: World Health Organization

## Code availability

Cannot be applied.

## Author information

### Authors and Affiliations

**FSBI “Federal Center of Neurosurgery”, Nemirovich-Danchenko Str. 132/1, Novosibirsk 630087, Russia**

Elena Filimonova, Anton Pashkov, Anton Kalinovsky, Jamil Rzaev

**Department of Neurosurgery, Novosibirsk State Medical University, Krasny Prospect St. 52, Novosibirsk 630091, Russia**

Elena Filimonova, Norayr Borisov, Anton Kalinovsky, Jamil Rzaev

**Department of Data Collection and Processing Systems, Novosibirsk State Technical University, Karl Marx Avenue 20 Novosibirsk, Russia**

Anton Pashkov

**Department of Neuroscience, Institute of Medicine and Psychology, Novosibirsk State University, Pirogov Str. 1, Novosibirsk 630090, Russia**

Jamil Rzaev

### Corresponding author

Correspondence to Elena Filimonova

## Ethics declaration

### Ethical approval

Data acquisition and publication were in accordance with the principles outlined in the Declaration of Helsinki. The study was approved by the local Ethics Committee of the Federal Center for Neurosurgery, Novosibirsk, Russia (protocol No. 4 dated 02-08-2022).

### Informed consent

Informed consent was obtained from all individual participants included in the study.

### Consent for publication

All patients provided informed consent for publication.

## References

[1] A. Pellerino, M. Caccese, M. Padovan, G. Cerretti, and G. Lombardi, “Epidemiology, risk factors, and prognostic factors of gliomas,” Clin Transl Imaging, vol. 10, no. 5, pp. 467–475, Oct. 2022, doi: 10.1007/S40336-022-00489-6/METRICS.

[2] C. Horbinski et al., “NCCN Guidelines® Insights: Central Nervous System Cancers, Version 2.2022: Featured Updates to the NCCN Guidelines,” Journal of the National Comprehensive Cancer Network, vol. 21, no. 1, pp. 12–20, Jan. 2023, doi: 10.6004/JNCCN.2023.0002.

[3] B. Powter et al., “Human TERT promoter mutations as a prognostic biomarker in glioma,” J Cancer Res Clin Oncol, vol. 147, no. 4, pp. 1007–1017, Apr. 2021, doi: 10.1007/S00432-021-03536-3/TABLES/2.

[4] A. Wenger and H. Carén, “Methylation Profiling in Diffuse Gliomas: Diagnostic Value and Considerations,” Cancers (Basel), vol. 14, no. 22, Nov. 2022, doi: 10.3390/CANCERS14225679.

[5] D. N. Louis et al., “The 2021 WHO Classification of Tumors of the Central Nervous System: a summary,” Neuro Oncol, vol. 23, no. 8, pp. 1231–1251, Aug. 2021, doi: 10.1093/NEUONC/NOAB106.

[6] J. Ker, Y. Bai, H. Y. Lee, J. Rao, and L. Wang, “Automated brain histology classification using machine learning,” Journal of Clinical Neuroscience, vol. 66, pp. 239–245, Aug. 2019, doi: 10.1016/j.jocn.2019.05.019.

[7] M. Ideguchi et al., “Investigation of histological heterogeneity based on the discrepancy between the hyperintense area on T2-weighted images and the accumulation area on 11C-methionine PET in minimally enhancing glioma,” Interdisciplinary Neurosurgery, vol. 27, p. 101364, Mar. 2022, doi: 10.1016/J.INAT.2021.101364.

[8] L. A. G. Nielsen et al., “Evaluation of the proliferation marker Ki-67 in gliomas: Interobserver variability and digital quantification,” Diagn Pathol, vol. 13, no. 1, Jun. 2018, doi: 10.1186/S13000-018-0711-2.

[9] X. Wang et al., “Combining Radiology and Pathology for Automatic Glioma Classification,” Front Bioeng Biotechnol, vol. 10, p. 841958, Mar. 2022, doi: 10.3389/FBIOE.2022.841958/BIBTEX.

[10] S. S. Tebha, S. Ali Memon, Q. Mehmood, D. Mukherjee, H. Abdi, and A. Negida, “Glioblastoma management in low and middle-income countries; existing challenges and policy recommendations,” Brain and Spine, vol. 3, p. 101775, Jan. 2023, doi: 10.1016/J.BAS.2023.101775.

[11] N. Upadhyay and A. D. Waldman, “Conventional MRI evaluation of gliomas,” Br J Radiol, vol. 84, no. Spec Iss 2, p. S107, Dec. 2011, doi: 10.1259/BJR/65711810.

[12] Y. Li et al., “Radiomics-Based Method for Predicting the Glioma Subtype as Defined by Tumor Grade, IDH Mutation, and 1p/19q Codeletion,” Cancers 2022, Vol. 14, Page 1778, vol. 14, no. 7, p. 1778, Mar. 2022, doi: 10.3390/CANCERS14071778.

[13] W. You et al., “The combination of radiomics features and VASARI standard to predict glioma grade,” Front Oncol, vol. 13, p. 1083216, Mar. 2023, doi: 10.3389/FONC.2023.1083216/BIBTEX.

[14] N. Du et al., “Preoperative and Noninvasive Prediction of Gliomas Histopathological Grades and IDH Molecular Types Using Multiple MRI Characteristics,” Front Oncol, vol. 12, p. 873839, May 2022, doi: 10.3389/FONC.2022.873839/BIBTEX.

[15] N. Haydar et al., “Role of Magnetic Resonance Imaging (MRI) in grading gliomas comparable with pathology: A cross-sectional study from Syria,” Annals of Medicine and Surgery, vol. 82, p. 104679, Oct. 2022, doi: 10.1016/J.AMSU.2022.104679.

[16] A. Lasocki et al., “Conventional MRI features can predict the molecular subtype of adult grade 2–3 intracranial diffuse gliomas,” Neuroradiology, vol. 64, no. 12, pp. 2295–2305, Dec. 2022, doi: 10.1007/S00234-022-02975-0/FIGURES/3.

[17] L. Gemini et al., “Vasari Scoring System in Discerning between Different Degrees of Glioma and IDH Status Prediction: A Possible Machine Learning Application?,” J Imaging, vol. 9, no. 4, Apr. 2023, doi: 10.3390/JIMAGING9040075.

[18] J. J. M. Van Griethuysen et al., “Computational radiomics system to decode the radiographic phenotype,” Cancer Res, vol. 77, no. 21, pp. e104–e107, Nov. 2017, doi: 10.1158/0008-5472.CAN-17-0339/SUPPLEMENTARY-VIDEO-S2.

[19] M. W. Wagner et al., “Radiomics of Pediatric Low-Grade Gliomas: Toward a Pretherapeutic Differentiation of BRAF-Mutated and BRAF-Fused Tumors,” AJNR Am J Neuroradiol, vol. 42, no. 4, p. 759, Apr. 2021, doi: 10.3174/AJNR.A6998.

[20] Y. Su et al., “Whole-tumor histogram analysis of diffusion and perfusion metrics for noninvasive pediatric glioma grading,” Neuroradiology, vol. 65, no. 6, pp. 1063–1071, Jun. 2023, doi: 10.1007/S00234-023-03145-6.

[21] N. C. Nuessle et al., “ADC-Based Stratification of Molecular Glioma Subtypes Using High b-Value Diffusion-Weighted Imaging,” J Clin Med, vol. 10, no. 16, p. 3451, Aug. 2021, doi: 10.3390/JCM10163451.

[22] N. Du et al., “An initial study on the predictive value using multiple MRI characteristics for Ki-67 labeling index in glioma,” J Transl Med, vol. 21, no. 1, pp. 1–11, Dec. 2023, doi: 10.1186/S12967-023-03950-W/FIGURES/5.

[23] R. Yao, A. Cheng, Z. Zhang, B. Jin, and H. Yu, “Correlation Between Apparent Diffusion Coefficient and the Ki-67 Proliferation Index in Grading Pediatric Glioma,” J Comput Assist Tomogr, vol. 47, no. 2, p. 322, Mar. 2023, doi: 10.1097/RCT.0000000000001400.

[24] A. Hilario et al., “The added value of apparent diffusion coefficient to cerebral blood volume in the preoperative grading of diffuse gliomas,” AJNR Am J Neuroradiol, vol. 33, no. 4, pp. 701–707, Apr. 2012, doi: 10.3174/AJNR.A2846.

[25] W. Phuttharak, J. Thammaroj, S. Wara-Asawapati, and K. Panpeng, “Grading Gliomas Capability: Comparison between Visual Assessment and Apparent Diffusion Coefficient (ADC) Value Measurement on Diffusion-Weighted Imaging (DWI),” Asian Pac J Cancer Prev, vol. 21, no. 2, p. 385, Feb. 2020, doi: 10.31557/APJCP.2020.21.2.385.

[26] R. K. Soliman, A. A. Essa, A. A. S. Elhakeem, S. A. Gamal, and M. M. A. Zaitoun, “Texture analysis of apparent diffusion coefficient (ADC) map for glioma grading: Analysis of whole tumoral and peri-tumoral tissue,” Diagn Interv Imaging, vol. 102, no. 5, pp. 287–295, May 2021, doi: 10.1016/J.DIII.2020.12.001.

[27] T. B. Nguyen et al., “Correlation of Tumor Immunohistochemistry with Dynamic Contrast-Enhanced and DSC-MRI Parameters in Patients with Gliomas,” AJNR Am J Neuroradiol, vol. 37, no. 12, p. 2217, Dec. 2016, doi: 10.3174/AJNR.A4908.

[28] J. Liang et al., “Diagnostic Values of DCE-MRI and DSC-MRI for Differentiation Between High-grade and Low-grade Gliomas: A Comprehensive Meta-analysis,” Acad Radiol, vol. 25, no. 3, pp. 338–348, Mar. 2018, doi: 10.1016/J.ACRA.2017.10.001.

[29] L. Siakallis, C. C. Topriceanu, J. Panovska-Griffiths, and S. Bisdas, “The role of DSC MR perfusion in predicting IDH mutation and 1p19q codeletion status in gliomas: meta-analysis and technical considerations,” Neuroradiology, vol. 65, no. 7, pp. 1111–1126, Jul. 2023, doi: 10.1007/S00234-023-03154-5/FIGURES/5.

[30] M. A. Schmidt et al., “Standardized acquisition and post-processing of dynamic susceptibility contrast perfusion in patients with brain tumors, cerebrovascular disease and dementia: comparability of post-processing software,” Br J Radiol, vol. 93, no. 1105, 2020, doi: 10.1259/BJR.20190543.

[31] M. Grade, J. A. Hernandez Tamames, F. B. Pizzini, E. Achten, X. Golay, and M. Smits, “A neuroradiologist’s guide to arterial spin labeling MRI in clinical practice,” Neuroradiology, vol. 57, no. 12, p. 1181, Dec. 2015, doi: 10.1007/S00234-015-1571-Z.

[32] A. Alsaedi et al., “The value of arterial spin labelling in adults glioma grading: systematic review and meta-analysis,” Oncotarget, vol. 10, no. 16, p. 1589, Feb. 2019, doi: 10.18632/ONCOTARGET.26674.

[33] N. Wang, S. yi Xie, H. ming Liu, G. quan Chen, and W. dong Zhang, “Arterial Spin Labeling for Glioma Grade Discrimination: Correlations with IDH1 Genotype and 1p/19q Status,” Transl Oncol, vol. 12, no. 5, p. 749, May 2019, doi: 10.1016/J.TRANON.2019.02.013.

[34] A. A. ElBeheiry, D. M. Emara, A. A. B. Abdel-Latif, M. Abbas, and A. S. Ismail, “Arterial spin labeling in the grading of brain gliomas: could it help?,” Egyptian Journal of Radiology and Nuclear Medicine, vol. 51, no. 1, pp. 1–11, Dec. 2020, doi: 10.1186/S43055-020-00352-6/TABLES/6.

[35] T. Hashido, S. Saito, and T. Ishida, “A radiomics-based comparative study on arterial spin labeling and dynamic susceptibility contrast perfusion-weighted imaging in gliomas,” Scientific Reports 2020 10:1, vol. 10, no. 1, pp. 1–10, Apr. 2020, doi: 10.1038/s41598-020-62658-9.

[36] Z. Wen et al., “MR imaging of high-grade brain tumors using endogenous protein and peptide-based contrast,” Neuroimage, vol. 51, no. 2, pp. 616–622, Jun. 2010, doi: 10.1016/J.NEUROIMAGE.2010.02.050.

[37] C. K. Jones, M. J. Schlosser, P. C. M. Van Zijl, M. G. Pomper, X. Golay, and J. Zhou, “Amide proton transfer imaging of human brain tumors at 3T,” Magn Reson Med, vol. 56, no. 3, pp. 585–592, 2006, doi: 10.1002/MRM.20989.

[38] B. Sotirios, E. Demetriou, C. C. Topriceanu, and Z. Zakrzewska, “The role of APT imaging in gliomas grading: A systematic review and meta-analysis,” Eur J Radiol, vol. 133, p. 109353, Dec. 2020, doi: 10.1016/J.EJRAD.2020.109353.

[39] S. Jiang et al., “Amide proton transfer-weighted magnetic resonance image-guided stereotactic biopsy in patients with newly diagnosed gliomas,” Eur J Cancer, vol. 83, pp. 9–18, Sep. 2017, doi: 10.1016/J.EJCA.2017.06.009.

[40] J. Zhou, H. Y. Heo, L. Knutsson, P. C. M. van Zijl, and S. Jiang, “APT-weighted MRI: Techniques, current neuro applications, and challenging issues,” Journal of Magnetic Resonance Imaging, vol. 50, no. 2, pp. 347–364, Aug. 2019, doi: 10.1002/JMRI.26645.

[41] A. I. Friismose, L. Markovic, N. Nguyen, O. Gerke, M. K. Schulz, and B. R. Mussmann, “Amide proton transfer-weighted MRI in the clinical setting – correlation with dynamic susceptibility contrast perfusion in the post-treatment imaging of adult glioma patients at 3T,” Radiography, vol. 28, no. 1, pp. 95–101, Feb. 2022, doi: 10.1016/J.RADI.2021.08.006.

[42] X. W. Kang et al., “Grading of Glioma: Combined diagnostic value of amide proton transfer weighted, arterial spin labeling and diffusion weighted magnetic resonance imaging,” BMC Med Imaging, vol. 20, no. 1, pp. 1–8, May 2020, doi: 10.1186/S12880-020-00450-X/FIGURES/2.

[43] M. Nakajo, M. Bohara, K. Kamimura, N. Higa, and T. Yoshiura, “Correlation between amide proton transfer-related signal intensity and diffusion and perfusion magnetic resonance imaging parameters in high-grade glioma,” Scientific Reports 2021 11:1, vol. 11, no. 1, pp. 1–7, May 2021, doi: 10.1038/s41598-021-90841-z.

[44] Y. Bai et al., “Noninvasive amide proton transfer magnetic resonance imaging in evaluating the grading and cellularity of gliomas,” Oncotarget, vol. 8, no. 4, pp. 5834–5842, 2017, doi: 10.18632/ONCOTARGET.13970.

[45] Z. Xu et al., “Diagnostic performance between MR amide proton transfer (APT) and diffusion kurtosis imaging (DKI) in glioma grading and IDH mutation status prediction at 3 T,” Eur J Radiol, vol. 134, Jan. 2021, doi: 10.1016/J.EJRAD.2020.109466.

[46] H. Guo et al., “Diagnostic performance of gliomas grading and IDH status decoding A comparison between 3D amide proton transfer APT and four diffusion-weighted MRI models,” Journal of Magnetic Resonance Imaging, vol. 56, no. 6, pp. 1834–1844, Dec. 2022, doi: 10.1002/JMRI.28211.

[47] B. Joo et al., “Amide proton transfer imaging might predict survival and IDH mutation status in high-grade glioma,” Eur Radiol, vol. 29, no. 12, p. 6643, Dec. 2019, doi: 10.1007/S00330-019-06203-X.

[48] K. Chen, X. W. Jiang, L. J. Deng, and H. L. She, “Differentiation between glioma recurrence and treatment effects using amide proton transfer imaging: A mini-Bayesian bivariate meta-analysis,” Front Oncol, vol. 12, p. 852076, Aug. 2022, doi: 10.3389/FONC.2022.852076/BIBTEX.

[49] N. von Knebel Doeberitz et al., “CEST imaging of the APT and ssMT predict the overall survival of patients with glioma at the first follow-up after completion of radiotherapy at 3T,” Radiother Oncol, vol. 184, Jul. 2023, doi: 10.1016/J.RADONC.2023.109694.

[50] Y. S. Choi et al., “Amide proton transfer imaging to discriminate between low- and high-grade gliomas: added value to apparent diffusion coefficient and relative cerebral blood volume,” Eur Radiol, vol. 27, no. 8, pp. 3181–3189, Aug. 2017, doi: 10.1007/S00330-017-4732-0/FIGURES/5.

[51] N. Salari, R. Fatahian, M. Kazeminia, A. Hosseinian-Far, S. Shohaimi, and M. Mohammadi, “Patients’ Survival with Astrocytoma After Treatment: a Systematic Review and Meta-analysis of Clinical Trial Studies,” Indian J Surg Oncol, vol. 13, no. 2, p. 329, Jun. 2022, doi: 10.1007/S13193-022-01533-7.

[52] J. S. Marra, G. P. Mendes, G. H. Yoshinari, F. da Silva Guimarães, S. C. Mazin, and H. F. de Oliveira, “Survival after radiation therapy for high-grade glioma,” Reports of Practical Oncology & Radiotherapy, vol. 24, no. 1, pp. 35–40, Jan. 2019, doi: 10.1016/J.RPOR.2018.09.003.

[53] D. N. Yeboa et al., “Differences in patterns of care and outcomes between grade II and grade III molecularly defined 1p19q co-deleted gliomas,” Clin Transl Radiat Oncol, vol. 15, pp. 46– 52, Feb. 2019, doi: 10.1016/J.CTRO.2018.12.003.

[54] Y. Han et al., “Amide Proton Transfer Imaging in Predicting Isocitrate Dehydrogenase 1 Mutation Status of Grade II/III Gliomas Based on Support Vector Machine,” Front Neurosci, vol. 14, p. 510647, Feb. 2020, doi: 10.3389/FNINS.2020.00144/BIBTEX.

[55] C. Su et al., “Multi-parametric Z-spectral MRI may have a good performance for glioma stratification in clinical patients,” Eur Radiol, vol. 32, no. 1, pp. 101–111, Jan. 2022, doi: 10.1007/S00330-021-08175-3/METRICS.

[56] S. Qu, O. Qiu, and Z. Hu, “The prognostic factors and nomogram for patients with high-grade gliomas,” Fundamental Research, vol. 1, no. 6, pp. 824–828, Nov. 2021, doi: 10.1016/J.FMRE.2021.07.005.

[57] Z. Jia et al., “Exploring the relationship between age and prognosis in glioma: rethinking current age stratification,” BMC Neurol, vol. 22, no. 1, pp. 74–88, Dec. 2022, doi: 10.1186/S12883-022-02879-9.

[58] Z. Lin et al., “Establishment of age group classification for risk stratification in glioma patients,” BMC Neurol, vol. 20, no. 1, Aug. 2020, doi: 10.1186/S12883-020-01888-W.

[59] O. Togao et al., “Grading diffuse gliomas without intense contrast enhancement by amide proton transfer MR imaging: comparisons with diffusion- and perfusion-weighted imaging,” Eur Radiol, vol. 27, no. 2, pp. 578–588, Feb. 2017, doi: 10.1007/S00330-016-4328-0.

[60] C. Brendle et al., “Glioma Grading and Determination of IDH Mutation Status and ATRX loss by DCE and ASL Perfusion,” Clin Neuroradiol, vol. 28, no. 3, pp. 421–428, Sep. 2018, doi: 10.1007/S00062-017-0590-Z.

[61] Y. Yin et al., “Correlation of apparent diffusion coefficient with Ki-67 in the diagnosis of gliomas,” Zhongguo Yi Xue Ke Xue Yuan Xue Bao, vol. 34, no. 5, pp. 503–508, Oct. 2012, doi: 10.3881/J.ISSN.1000-503X.2012.05.012.

[62] C. Su et al., “Amide Proton Transfer Imaging Allows Detection of Glioma Grades and Tumor Proliferation: Comparison with Ki-67 Expression and Proton MR Spectroscopy Imaging,” AJNR Am J Neuroradiol, vol. 38, no. 9, p. 1702, Sep. 2017, doi: 10.3174/AJNR.A5301.

[63] O. Togao et al., “Amide proton transfer imaging of adult diffuse gliomas: correlation with histopathological grades,” Neuro Oncol, vol. 16, no. 3, p. 441, Mar. 2014, doi: 10.1093/NEUONC/NOT158.

[64] Y. Bai et al., “Noninvasive amide proton transfer magnetic resonance imaging in evaluating the grading and cellularity of gliomas,” Oncotarget, vol. 8, no. 4, pp. 5834–5842, 2017, doi: 10.18632/ONCOTARGET.13970.

