## Supplementary material for "Utilizing the Amide Proton Transfer Technique to Characterize Diffuse Gliomas Based on the WHO 2021 Classification of CNS Tumors": Table 1

***Table 1.*** *Parameters of MRI sequences*

|  | ***T1WI/CE-T1WI*** | ***T2WI*** | ***FLAIR*** | ***APT*** | ***ASL*** | ***ADC*** |
| --- | --- | --- | --- | --- | --- | --- |
| *Sequence type* | *3D TFE* | *3D TSE* | *3D TSE* | *3D SE* | *3D pCASL* | *2D EPI* |
| *TR (msec)* | *6.7* | *3500* | *4800* | *5925* | *4300* | *3800* |
| *TE (msec)* | *2.98* | *300* | *340* | *8.3* | *11.6* | *90* |
| *Flip angle* | *8* | *90* | *90* | *90* | *90* | *90* |
| *Matrix* | *256х256* | *256х256* | *228x228* | *128x128* | *64x64* | *128x128* |
| *FOV (mm)* | *256* | *256* | *256* | *256* | *240* | *256* |
| *Slice thickness (mm)* | *1* | *1* | *1.2* | *6* | *6* | *4* |
| *N of slices* | *192* | *360* | *140* | *10* | *14* | *26* |
| *Inversion time (msec)* | */* | */* | *1650* | */* | */* | */* |
| *b values* | */* | */* | */* | */* | */* | *0, 500, 1000* |
| *PLD (msec)* | */* | */* | */* | */* | *1800* | */* |
