## Supplementary material for "Utilizing the Amide Proton Transfer Technique to Characterize Diffuse Gliomas Based on the WHO 2021 Classification of CNS Tumors": Table 2

|  | **Third grade** | | | **Fourth grade** | | |
| --- | --- | --- | --- | --- | --- | --- |
| **Characteristic** | **OR** | **95% CI** | **p-value** | **OR** | **95% CI** | **p-value** |
| Median APT | 1.33 | 0.24, 7.32 | 0.7 | 8.52 | 1.28, 56.5 | 0.026* |
| Age | 0.96 | 0.89, 1.04 | 0.3 | 1.05 | 0.97, 1.15 | 0.2 |

**Table 2.** Parameters of the best (two-predictors) model, classifying patients into three tumor grades. Reference category was chosen to be the second grade. Table provides the odds ratio, 95% confidence interval and corresponding p-values for each predictor in the model.  OR - odds ratio, CI - confidence interval. * statistical significance at the level of p<0.05
