## Supplementary material for "Utilizing the Amide Proton Transfer Technique to Characterize Diffuse Gliomas Based on the WHO 2021 Classification of CNS Tumors": Table 3

|  | **Glioblastoma** | | | **Oligodendroglioma** | | |
| --- | --- | --- | --- | --- | --- | --- |
| **Characteristic** | **OR** | **95% CI** | **p-value** | **OR** | **95% CI** | **p-value** |
| Median APT | 203 | 16.5, 249.7 | 0.03* | 0.36 | 0.07, 1.83 | 0.2 |
| Age | 1.21 | 1.04, 1.41 | 0.013* | 1.03 | 0.96, 1.11 | 0.4 |
| necrosis | 7.04 | 0.70, 70.9 | 0.1 | 0.37 | 0.12, 1.19 | 0.095 |
| hemorrage | 0.01 | 0.00, 7.34 | 0.2 | 19.4 | 1.54, 243 | 0.022* |

**Table 3.** Parameters of the best (four-predictors) model, classifying patients into three tumor types. Reference category was chosen to be the Astrocytoma group. Table provides the odds ratio, 95% confidence interval and corresponding p-values for each predictor in the model.  OR - odds ratio, CI - confidence interval. * statistical significance at the level of p<0.05
