## Supplementary material for "Utilizing the Amide Proton Transfer Technique to Characterize Diffuse Gliomas Based on the WHO 2021 Classification of CNS Tumors": Table 4

| **Characteristic** | **OR** | **95% CI** | **p-value** |
| --- | --- | --- | --- |
| Median APT | 32.5 | 4.78, 626 | 0.004** |
| Age | 1.16 | 1.06, 1.33 | 0.009** |

**Table 4.** Parameters of the best (two-predictors) model, classifying patients into two IDH1 groups. Table provides the odds ratio, 95% confidence interval and corresponding p-values for each predictor included in the model.  Mutant type was chosen to be a reference level. OR - odds ratio, CI - confidence interval. ** statistical significance at the level of p<0.01
