## Supplementary Table 3 for "Utilizing the Amide Proton Transfer Technique to Characterize Diffuse Gliomas Based on the WHO 2021 Classification of CNS Tumors"

**Mean APT values between Grade groups**

```
Kruskal-Wallis chi-squared = 8.302, df = 2, p-value = 0.016

Dunn (1964) Kruskal-Wallis multiple comparison
  p-values adjusted with the Benjamini-Hochberg method.

Comparison          Z    P.unadj      P.adj
1      1 - 2 -0.4456362 0.65586008 0.65586008
2      1 - 3 -2.1500656 0.03155002 0.04732503
3      2 - 3 -2.3882969 0.01692666 0.05077998
```

**Mean APT values between tumor type groups**

```
Kruskal-Wallis chi-squared = 13.026, df = 2, p-value = 0.001484

Dunn (1964) Kruskal-Wallis multiple comparison
  p-values adjusted with the Benjamini-Hochberg method.

                        Comparison         Z      P.unadj       P.adj
1       astrocytoma - glioblastoma -2.769439 0.0056152859 0.008422929
2  astrocytoma - olygodendroglioma  1.053801 0.2919741811 0.291974181
3 glioblastoma - olygodendroglioma  3.316636 0.0009110826 0.002733248
```

**Mean APT values between IDH1 status groups**

Wilcoxon rank sum exact test

W = 71, p-value = 0.0003306

**Mean APT values between 1p19q status groups**

Wilcoxon rank sum exact test

W = 224, p-value = 0.01957

**90^th^ percentile APT values between Grade groups**

```
Kruskal-Wallis chi-squared = 10.828, df = 2, p-value = 0.004454

Dunn (1964) Kruskal-Wallis multiple comparison
  p-values adjusted with the Benjamini-Hochberg method.

  Comparison          Z     P.unadj      P.adj
1      1 - 2 -0.9460832 0.344106157 0.34410616
2      1 - 3 -2.7304176 0.006325415 0.01897625
3      2 - 3 -2.4520253 0.014205468 0.02130820
```

**90^th^ percentile APT values between tumor type groups**

```
Kruskal-Wallis chi-squared = 13.266, df = 2, p-value = 0.001316

Dunn (1964) Kruskal-Wallis multiple comparison
  p-values adjusted with the Benjamini-Hochberg method.

                        Comparison          Z     P.unadj       P.adj
1       astrocytoma - glioblastoma -3.1347255 0.001720150 0.005160449
2  astrocytoma - olygodendroglioma  0.4880761 0.625495935 0.625495935
3 glioblastoma - olygodendroglioma  3.0717548 0.002128045 0.003192067
```

**90^th^ percentile APT values between IDH1 status groups**

Wilcoxon rank sum exact test

W = 65, p-value = 0.0001576

**90^th^ percentile APT values between 1p19q status groups**

Wilcoxon rank sum exact test

W = 209, p-value = 0.06523

**Mean ADC values between Grade groups**

Kruskal-Wallis chi-squared = 6.7128, df = 2, p-value = 0.03486

```
Dunn (1964) Kruskal-Wallis multiple comparison
  p-values adjusted with the Benjamini-Hochberg method.

  Comparison         Z    P.unadj     P.adj
1      1 - 2 0.5028301 0.61508372 0.6150837
2      1 - 3 2.0008323 0.04541047 0.0681157
3      2 - 3 2.0878631 0.03681018 0.1104306
```

**Mean ADC values between tumor type groups**

Kruskal-Wallis chi-squared = 3.1395, df = 2, p-value = 0.2081

**Mean ADC values between IDH1 status groups**

Wilcoxon rank sum exact test

W = 270, p-value = 0.0785

**Mean ADC values between 1p19q status groups**

Wilcoxon rank sum exact test

W = 126, p-value = 0.5066

**10^th^ percentile ADC values between Grade groups**

```
Kruskal-Wallis chi-squared = 6.3599, df = 2, p-value = 0.04159

Dunn (1964) Kruskal-Wallis multiple comparison
  p-values adjusted with the Benjamini-Hochberg method.

  Comparison        Z    P.unadj     P.adj
1      1 - 2 0.636360 0.52454181 0.5245418
2      1 - 3 2.039770 0.04137320 0.1241196
3      2 - 3 1.939399 0.05245274 0.0786791
```

**10^th^ percentile ADC values between tumor type groups**

Kruskal-Wallis chi-squared = 3.614, df = 2, p-value = 0.1641

**10^th^ percentile ADC values between IDH1 status groups**

Wilcoxon rank sum test with continuity correction

W = 268.5, p-value = 0.08551

**10^th^ percentile ADC values between 1p19q status groups**

Wilcoxon rank sum test with continuity correction

W = 150, p-value = 0.9755
