## Supplementary Table 1 for "Utilizing the Amide Proton Transfer Technique to Characterize Diffuse Gliomas Based on the WHO 2021 Classification of CNS Tumors"

***Supplementary table 1.*** *The detailed information about patients.*

|  | ***Gender*** | ***Age*** | ***Tumor location*** | ***Tumor type*** | ***Ki67, %*** | ***Grade*** | ***IDH1*** | ***1p19q*** |
| --- | --- | --- | --- | --- | --- | --- | --- | --- |
| *1* | male | 31-35 | right temporal lobe | astrocytoma | 5 | 2 | mutant | no |
| *2* | male | 41-45 | left temporal lobe | astrocytoma | 4 | 2 | mutant | no |
| *3* | male | 66-70 | right frontal lobe | oligodendroglioma | 4 | 2 | mutant | yes |
| *4* | male | 26-30 | right frontal lobe | astrocytoma | 7 | 3 | mutant | no |
| *5* | female | 46-50 | right insula | astrocytoma | 7 | 3 | mutant | no |
| *6* | female | 36-40 | right temporal lobe | astrocytoma | 7 | 3 | mutant | no |
| *7* | male | 31-35 | left temporal lobe | oligodendroglioma | 8 | 3 | mutant | yes |
| *8* | female | 66-70 | right temporal lobe | oligodendroglioma | 35 | 3 | mutant | yes |
| *9* | male | 36-40 | right frontal lobe | astrocytoma | 28 | 4 | mutant | no |
| *10* | male | 66-70 | left insula | astrocytoma | 18 | 4 | mutant | no |
| *11* | male | 61-65 | right parietal lobe | astrocytoma | 35 | 4 | mutant | no |
| *12* | male | 51-55 | left temporal lobe | astrocytoma | 18 | 4 | mutant | no |
| *13* | female | 36-40 | right parietal lobe | astrocytoma | 40 | 4 | mutant | no |
| *14* | female | 66-70 | left parietal lobe | astrocytoma | 83 | 4 | mutant | no |
| *15* | female | 56-60 | corpus callosum | glioblastoma | 45 | 4 | wild | no |
| *16* | female | 76-80 | left parietal lobe | glioblastoma | 82 | 4 | wild | no |
| *17* | female | 66-70 | left frontal lobe | glioblastoma | 30 | 4 | wild | no |
| *18* | female | 51-55 | right frontal lobe | glioblastoma | 25 | 4 | wild | no |
| *19* | female | 71-75 | left frontal lobe | glioblastoma | 16 | 4 | wild | no |
| *20* | male | 66-70 | right temporal lobe | glioblastoma | 32 | 4 | wild | no |
| *21* | male | 41-45 | right frontal lobe | astrocytoma | 10 | 3 | mutant | no |
| *22* | male | 66-70 | right temporal lobe | astrocytoma | 19 | 4 | mutant | no |
| *23* | male | 41-45 | left parietal lobe | oligodendroglioma | 25 | 3 | mutant | yes |
| *24* | female | 26-30 | left frontal lobe | astrocytoma | 17 | 4 | mutant | no |
| *25* | male | 61-65 | left occipital lobe | oligodendroglioma | 19 | 3 | mutant | yes |
| *26* | female | 51-55 | left frontal lobe | astrocytoma | 7 | 3 | mutant | no |
| *27* | female | 31-35 | right frontal lobe | astrocytoma | 12 | 3 | mutant | no |
| *28* | female | 41-45 | right frontal lobe | oligodendroglioma | 60 | 3 | mutant | yes |
| *29* | male | 51-55 | right frontal lobe | oligodendroglioma | 24 | 3 | mutant | yes |
| *30* | female | 56-60 | right frontal lobe | oligodendroglioma | 5 | 2 | mutant | yes |
| *31* | female | 66-70 | left frontal lobe | glioblastoma | 24 | 4 | wild | no |
| *32* | male | 61-65 | right temporal lobe | astrocytoma | 19 | 4 | mutant | no |
| *33* | female | 71-75 | left parietal lobe | glioblastoma | 54 | 4 | wild | no |
| *34* | female | 51-54 | corpus callosum | glioblastoma | 35 | 4 | wild | no |
| *35* | male | 46-50 | left temporal lobe | glioblastoma | 17 | 4 | wild | no |
| *36* | female | 51-55 | brainstem | astrocytoma | 4 | 2 | mutant | no |
| *37* | male | 66-70 | left temporal lobe | glioblastoma | 38 | 4 | wild | no |
| *38* | female | 46-50 | brainstem | glioblastoma | 69 | 4 | wild | no |
| *39* | female | 56-60 | right frontal lobe | glioblastoma | 20 | 4 | wild | no |
| *40* | female | 71-75 | right temporal lobe | glioblastoma | 24 | 4 | wild | no |
| *41* | female | 56-60 | left frontal lobe | glioblastoma | 42 | 4 | wild | no |
| *42* | male | 21-25 | right frontal lobe | astrocytoma | 14 | 3 | mutant | no |
